# Common Genetic Risk Factors in ASD and ADHD Co-occurring Families

**DOI:** 10.1101/2022.05.15.22275109

**Authors:** Anbo Zhou, Xiaolong Cao, Vaidhyanathan Mahaganapathy, Marco Azaro, Christine Gwin, Sherri Wilson, Steven Buyske, Christopher W. Bartlett, Judy F. Flax, Linda M. Brzustowicz, Jinchuan Xing

**Affiliations:** Department of Genetics, Rutgers, The State University of New Jersey, Piscataway, NJ, USA; Department of Statistics, Rutgers, The State University of New Jersey, Piscataway, NJ, USA; The Steve Cindy Rasmussen Institute for Genomic Medicine, Battelle Center for Computational Biology, Abigail Wexner Research Institute at Nationwide Children’s Hospital; Department of Pediatrics, College of Medicine, The Ohio State University, Columbus, Ohio, USA; The Human Genetics Institute of New Jersey, Rutgers, The State University of New Jersey, Piscataway, NJ, USA; Division of Laboratory Medicine, Zhujiang Hospital, Southern Medical University, Guangzhou, China

## Abstract

Autism spectrum disorder (ASD) and attention-deficit/hyperactivity disorder (ADHD) are two major neurodevelopmental disorders that frequently co-occur. However, the genetic mechanism of the co-occurrence remains unclear. The New Jersey Language and Autism Genetics Study (NJLAGS) collected more than 100 families with at least one member affected by ASD. NJLAGS families show a high prevalence of ADHD and provide a good opportunity to study shared genetic risk factors for ASD and ADHD. The linkage study of the NJLAGS families revealed regions on chromosomes 12 and 17 that are significantly associated with ADHD. Using whole genome sequencing data on 272 samples from 73 NJLAGS families, we identified potential risk genes for ASD and ADHD. Within the linkage regions, we identified 36 genes that are associated with ADHD using a pedigree-based gene prioritization approach. *KDM6B* (Lysine Demethylase 6B) is the highest-ranking gene, which is a known risk gene for neurodevelopmental disorders, including ASD and ADHD. At the whole genome level, we identified 207 candidate genes from the analysis of both small variants and structure variants, including both known and novel genes. Using enrichment and protein-protein interaction network analyses, we identified gene ontology terms and pathways enriched for ASD and ADHD candidate genes, such as cilia function and cation channel activity. Candidate genes and pathways identified in our study provide a better understanding of the genetic etiology of ASD and ADHD and will lead to new diagnostic or therapeutic interventions for ASD and ADHD in the future.

## Introduction

Autism spectrum disorder (ASD) and attention-deficit/hyperactivity disorder (ADHD) are two neurodevelopmental disorders (NDDs) of high prevalence and severity. ASD is characterized by deficits in social interaction and social communication, and by restricted, repetitive, and stereotyped patterns of behavior, interests, and activities (1). ADHD is characterized by inattention and hyperactive/ impulsive symptoms (1). The two disorders have shown a high comorbid frequency where 20-50% of children with ADHD meet the criteria for ASD and 30-80% of ASD children meet the criteria for ADHD (2). Both conditions can have a severe negative impact on patients’ quality of life, and only worsen when co-occurring.

On the behavioral level, and especially regarding diagnosis and clinical care, it is often difficult to distinguish some of the behavioral characteristics that define ASD to those that define ADHD. Individual with ASD often suffer from symptoms consistent with ADHD. Lack of focus, impulsivity, and emotional outbursts may be present in both disorders yet the approach to intervention, both behaviorally and pharmacologically, can be substantially different. Clarifying the genetic relationship between these two disorders could inform future treatment decisions. If ASD and ADHD represent pleiotropic manifestations of the same disorder, a unified treatment approach might be effective to target the symptoms of both. If they stem from different etiologies, separate treatments for each might prove more effective. Therefore, a deep understanding of the genetic etiology of familial co-occurrence of ASD and ADHD is important to enhance treatment planning.

Extensive research has been conducted on both disorders and demonstrates overlapping genetic factors between the two conditions (1–7). For example, ADHD candidate causal genes *DRD3* and *MAOA* are cautiously positively associated with ASD (2). In genome-wide association studies (GWAS), 16 single nucleotide variants (SNVs) related to ADHD were found to be possibly involved in ASD and 25 SNVs related to ASD are possibly involved in ADHD (2). In copy number variation (CNV) studies, CNVs segregated with ADHD were also found to be enriched for ASD candidate genes. Correspondingly, ASD patients’ family members also carry ADHD diagnosed CNVs (8, 9). While the studies have suggested shared genetic risk factors as one possible reason for the co-occurrence of the two disorders, they either used microarray or whole exome sequencing data. In addition, those studies are based on the case-control study design and do not leverage the power of family studies. Therefore, the evidence of shared genetic risk factors between ASD and ADHD from these studies is mostly weak and inconclusive.

To further explore the genetic etiology of ASD and ADHD co-occurrence, we aimed to use whole-genome sequencing (10) analysis to identify SNVs and structural variants (SVs) responsible for the ASD or ADHD phenotypes segregated in the New Jersey Language and Autism Genetics Study (NJLAGS) project (11). NJLAGS is a project studying the genetic etiology of ASD and related NDDs and has conducted WGS on 272 samples from 73 families, each with at least one ASD patient. All family members have been characterized by extensive behavioral assessments of language and social functioning, restrictive and repetitive behaviors, and other co-occurring behaviors (11). Despite the original study’s focus on ASD, these families exhibit elevated rates of ADHD, with more than 40% of the ASD patients also diagnosed with ADHD. The co-occurrence of ASD and ADHD in the NJLAGS families provides an opportunity to identify genetic factors underlying the co-occurring ASD and ADHD.

## Patients and Methods

### Sample collection and phenotype assessment

The sample in this study includes 79 families from a previous NJLAGS study (11) (Wave 1) plus an additional 32 families collected after the 2014 publication (Wave 2). The original aim of the NJLAGS study was to identify genetic variation related to DSM-IV autistic disorder, language impairment, as well as related disorders. Families were required to have at least one person with autistic disorder and one additional person with a language-learning impairment (11). During Wave 1, autism probands were required to have a strict diagnosis of autistic disorder based on the Autism Diagnostic Interview (ADI-R), Autism Diagnostic Observation Schedule (ADOS), and the Diagnostics and Statistical Manual-IV (DSM-IV). A second proband had to meet the criteria for Specific Language Impairment (SLI), a disorder where language development is delayed or deviant and cannot be explained by any other neurodevelopmental diagnosis. For Wave 2, NJLAGS sought to understand if the original strict autistic disorder diagnostic criteria were necessary, and therefore, the autism proband requirements were intentionally relaxed to become more in line with the less restrictive, newer DSM-5 criteria for Autism Spectrum Disorder (12). All subjects gave informed consent or assent conforming to the guidelines for treatment of human subjects from the Institutional Review Board at Rutgers, The State University of New Jersey (IRB number: 13-112Mc).

For ASD probands, ADHD characteristics were identified during the intake interview by either a study child psychiatrist or a developmental pediatrician. In each case, the diagnosis was corroborated in detailed NJLAGS history questionnaires that asked the parents if their child had ever received a diagnosis of attention deficit disorder (ADD) and/or ADHD: 1) Medical History Questionnaire, 2) Family History Questionnaire, and 3) Language Correlates Questionnaire. For all other family members, parents completed the same detailed Family History Questionnaire and Language Correlates Questionnaire regarding whether they or their children have received a diagnosis of several different neurodevelopmental or neuropsychiatric disorders, including ADD/ADHD. In all questionnaires ADD and ADHD were treated as one category and we used ADHD as a collective term for the affected individual.

### Genotyping and sequencing

DNA extraction was performed by RUCDR (Piscataway, NJ), either from blood DNA (WB) or Lymphoblastoid cell lines (LCL). Wave 1 Affymetrix Axiom array genotyping experiments have been described previously (11). For Wave 2, single nucleotide polymorphism (SNP) data was generated with the Illumina Infinium PsychArray-24 v1 array (Illumina, San Diego, CA), which includes 593,260 SNPs. SNP with a population minor allele frequency (MAF) >1% were selected. Quality control on SNP genotypes was conducted by array batch and by array type, as described previously (11). The quality control criteria include individual/SNP genotype completion, relationship checking, Mendelian errors checking, and ancestry inference. The linkage analysis only included samples that clustered with the CEU (Utah residents with Northern and Western European ancestry from the CEPH collection) samples from the HapMap reference data, as determined by EIGENSTRAT using the recommend parameters in the documentation (13).

Initially, a subset of 10,899 SNPs in common across the Wave 1 and 2 arrays was selected for the linkage analysis, which minimize marker-to-marker linkage disequilibrium while retaining a high MAF (>30%) to provide suitable genomic coverage of recombination events in the pedigrees. Overlap in some genomic regions was too low to retain acceptable information content as measured by MERLIN (14). In those regions, array-specific SNPs were included. This procedure did not pose an issue with missing data within families since every family was genotyped using only a single array type.

The whole-genome sequencing was performed in four batches by three vendors (Table S1). All samples were sequenced using Illumina paired-end format with a spec of 30x coverage of the human genome. Some samples were excluded in downstream analyses because of quality issues. A few were dropped because the subjects withdrew from the study. For samples that were sequenced in more than one batch, the best quality run was used for the analysis. The raw sequencing reads, variants, and genotypes for all samples are available in the National Institute of Mental Health Data Archive (NDA) under experiments C1932 and C2933.

### Linkage Analysis

Linkage analyses were conducted with KELVIN (v2.3.3). KELVIN implements the posterior probability of linkage (PPL) metric to estimate the probability that a genetic location is linked with a tested trait (15). Primary linkage analysis of the phenotypes was conducted on each wave separately, and the linkage evidence was sequentially updated across the waves using Bayes’ rule to provide a single metric for linkage evidence. A secondary linkage analysis was conducted using all families jointly in a single pooled analysis of each trait. By comparing the sequentially updated result to the pooled result, the role of heterogeneity in the dataset can be inferred qualitatively. Because stratifying on an irrelevant trait will on average produce the same result as a pooled analysis (16), if the sequentially updated PPLs were appreciably higher than pooled results, heterogeneity demarcated by wave is likely to present in the data.

Based on previous simulations of the null distribution in the NJLAGS sample when correcting for three phenotypes (11), a PPL of 0.32 or greater is consistent with a genome-wide error rate of p<0.001, a PPL of 0.26 corresponds to p<0.01, and a PPL of 0.11 corresponds to p<0.05. These threshold values are similar to previous studies of the false positive rate of the PPL after correction for testing multiple phenotypes (17, 18).

### Small variant (SNV/indel) and SV calling

Alignment of paired-end FASTQ files was performed using the BWA-MEM algorithm (v0.7.12) (19) to the Human Genome Reference Consortium Build 37 (hg19) using default parameters. The output was converted to BAM format using SAMtools view (v0.1.19) (20). BAM files from read alignment were then processed using the GATK v3.5.0 variant calling pipeline following the best practice recommendation for alignment processing and variant calling (21, 22). Starting from sorted and indexed individual BAM files, a series of GATK alignment processing procedures were conducted, including PCR duplicate removal and base quality score recalibration. SNVs and small insertion/deletions (indels) were called per individual using HaplotypeCaller before being jointly called by GenotypeGVCF. All samples from different sequencing batches were jointly called along with the 1000 Genomes project European ancestry samples (CEU, GBR, FIN) as controls to reduce the batch-effect for downstream analysis (23). After variant call, we employed variant quality score recalibration using VariantRecalibrator and ApplyRecalibration as outlined in the GATK protocol.

SV calling has been historically difficult for tools using a single algorithm (24). Therefore, an ensemble algorithm, MetaSV (25), along with its components (Breakseq2 (26), breakdancer (27), CNVnator (28), and Pindel (29)) were used for SV discovery. Local realignment by SPAdes (30) and AGE (31) were carried out to further improve breakpoint resolution. MetaSV then combined all produced evidence into a single call set. The outputs of different chromosomes from the same individual were then merged back to one file using VCFtools (32). Basic statistics for the SV calls were calculated by SURVIVOR (33) for quality control. SVs were then filtered to select SVs that had consensus calls from at least two SV callers in MetaSV.

### Variant annotation and selection

SNVs and indels were annotated by VAT in the VAAST package (v2.0.2) (34) and condensed into CDR files to represent one family per file by VST in the VAAST package. The variants were filtered to include those that have an MAF < 5% in the ExAC dataset excluding psychiatric cohorts (ExAC non-psych v0.3.1 https://gnomad.broadinstitute.org/). For control samples, 635 GTEx WGS samples were obtained from the GTEx project (dbGaP accession number phs000424.v7.p2) (35) and condensed into a single group. SV calls were annotated by AnnotSV (v3.0.9) (36).

### Gene prioritization

For SNVs and indels, the gene prioritization tool pVAAST (v0.02) was used to prioritize candidate genes (37). A pVAAST score was calculated for each gene from its variants’ linkage pattern, association evidence, MAF, and functional prediction. pVAAST analyses were performed for the two ADHD linkage regions and the whole genome, under both dominant and recessive modes of inheritance. The linkage region analyses were performed at 10^6^ permutations per gene and the whole-genome analyses were performed at 10^5^ permutations per gene to determine the significance. For running pVAAST, the pedigrees were processed and trimmed to include individuals from one pair of ancestral parents per pedigree for the dominant mode, or a two-generation subset of the pedigree for the recessive mode. The individuals were selected to maximize the number of sequenced and affected samples in each family.

Candidate genes expressed in at least one of the brain expression databases were selected for downstream analysis. For linkage regions, candidate genes that had a Bonferroni-corrected p-value < 0.05 were considered significant and selected. For whole-genome analyses, candidate genes that have a p-value <= 10^−5^ and a LOD score >= 1 in the pVAAST analysis were selected for downstream analysis.

For SVs, AnnotSV annotation provides a ranking score for potential pathogenicity (36). SVs that are predicted to be likely pathogenic (class 4) or pathogenic (class 5) were selected. In each individual, the severity score of an SV in class 4 and 5 is calculated as: severity score = (class - 3) * number of SV copies, where the number of copies is set at 1 for heterozygous SVs, and 2 for homozygous SVs. A custom script was used to convert results from an SV-based report into a gene-based report. For each gene, the annotated severity scores of all SVs overlapping a given gene were aggregated for affected individuals and unaffected individuals separately. The genes were ranked based on the aggregated SV severity scores in affected individuals. Genes were selected for downstream analysis using the following criteria: 1. containing SV(s) in more than one affected individual; 2. expressed in at least one of the brain expression databases; 3. SV(s) overlapping the exonic region; and 4. the aggregated score in affected individuals is higher than that in unaffected individuals.

### Gene annotation, pathway and enrichment analyses

For gene function predictions, the pLI (probability of being loss-of-function intolerant) score was extracted from gnomAD (v3.0) for each gene (38). Brain developmental gene expression data were obtained from the GTEx project (v8) (35), the BrainSpan Atlas of the Developing Human Brain project (RNA-Seq GENCODE v10 summarized to genes, http://www.brainspan.org/static/download.html) (39), and the Human Developmental Biology Resource (https://www.ebi.ac.uk/gxa/experiments/E-MTAB-4840/Downloads) (40), as previously described (41). The genes’ association with specific cells or diseases are predicted by the HumanBase (https://hb.flatironinstitute.org) algorithm that learns from large genomics data collections (42). The gene knock-out mouse behavior was downloaded from the International Mouse Phenotyping Consortium (IMPC) (ftp://ftp.ebi.ac.uk/pub/databases/impc/latest/) (43). Candidate genes for several NDDs were collected from previous studies and the gene sets and genes are summarized in Table S2.

The protein-protein interaction (PPI) networks were generated based on gene interactions from three databases, including STRING (v11.5) (44), GIANT (v2.0) (42) and ConsensusPathDB (v35) (45). The cutoff for the “combined score” for STRING was set to 700. For GIANT, interactions with score > 0.6 in tissues from brain and neuron system were used, including brain, astrocyte_of_the_cerebellum, astrocyte_of_the_cerebral_cortex, basal_ganglion, brainstem, central_nervous_system, central_nervous_system_pericyte, cerebellar_cortex, cerebellum, cerebral_cortex, cerebral_hemisphere, corpus_striatum, forebrain, frontal_cortex, glial_cell, global, globus_pallidus, gray_matter, gray_matter_of_telencephalon, gyrus, hindbrain, hippocampal_formation, hypothalamus, lobe_of_cerebral_hemisphere, neocortex, nervous_system, neural_cell, neuron, nucleus_of_brain, pallium, parietal_lobe, peripheral_nervous_system, pituitary_gland, pons, spinal_cord, substantia_nigra, telencephalic_nucleus and telencephalon. The Python NetworkX package (https://networkx.org) was used to visualize the PPI network and the positions of genes in the network were saved for final plotting with R (v4.1.0) packages ggplot2 and scatterpie. Details of interaction data download and processing procedures were described previously (41, 46).

Gene set enrichment analyses were performed using over-representation analysis provided by ConsensusPathDB (45). All pathway, Gene Ontology (GO), and protein complex-based enrichment analyses were enabled, with the minimum two genes from the input and p-value cutoff set to 1. Enrichment p-values for all genes were determined by ConsensusPathDB using a hypergeometric test and enrichment p-value for each gene list was calculated with Fisher’s exact test as described previously (41). An enriched term (*i*.*e*., GO, pathway, or protein complex) was selected for further analysis if: 1) the total number of genes belong to the term is ≤ 500; 2) the term includes more than three genes from all input genes (ADHD, ASD/ADHD, and NDD genes); 3) enrichment p-value < 0.01 for ADHD and/or ASD/ADHD gene lists.

## Results

### Overall study design

For samples in the NJLAGS cohort, we conducted the SNP genotyping microarray analysis and the WGS analysis in parallel (Figure 1). Genotypes from the genotyping microarray were used to perform linkage analysis and the WGS data were then used to prioritize genes based on SNVs/indels and SVs in both linkage regions and the whole genome. To account for the genetic heterogeneity of ASD and ADHD, candidate gene sets were subjected to enrichment and pathway analyses.

**Figure 1.**
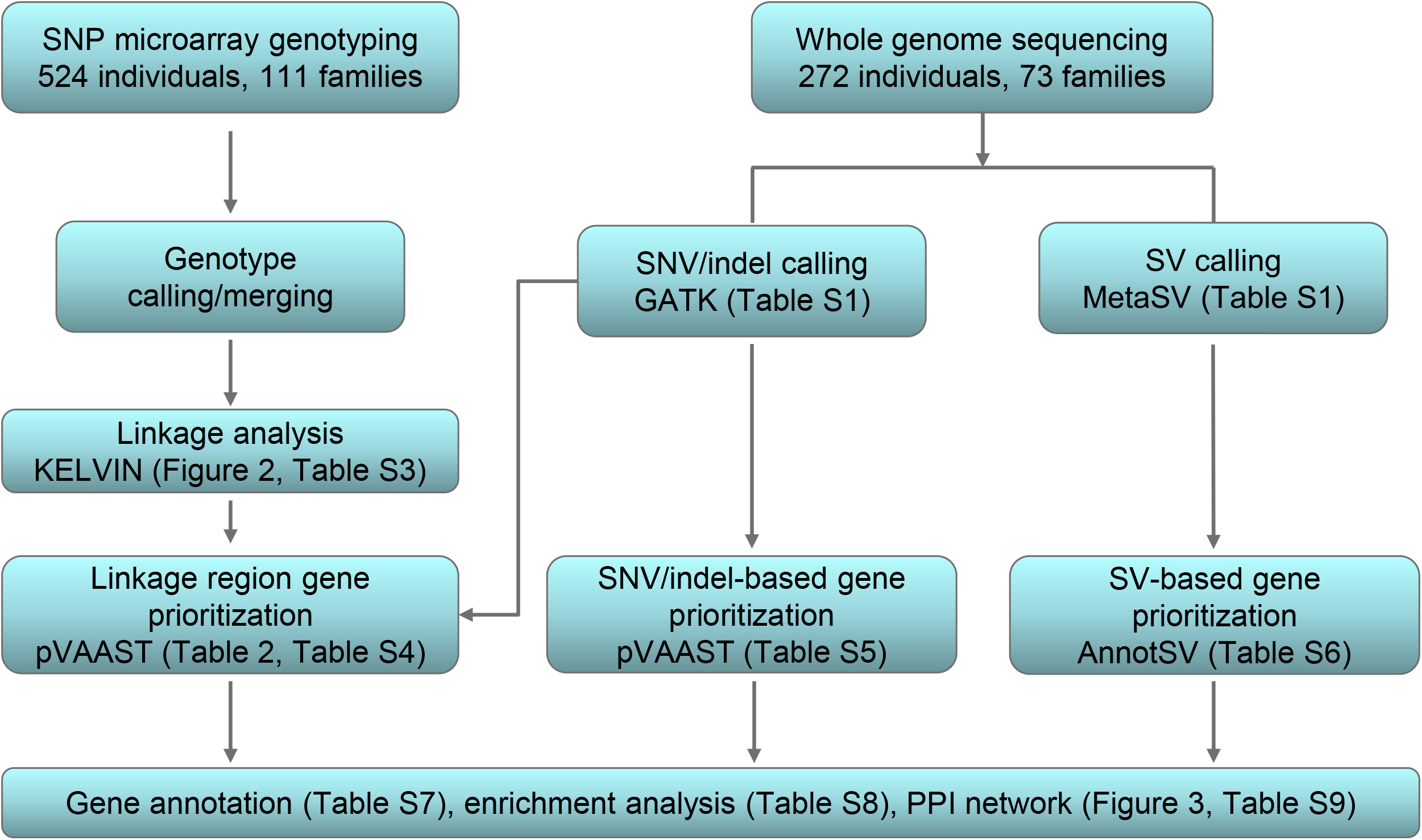
Project overview and overall analysis flow.

Among the 73 families involved in WGS, 47 (64.3%) have individuals who are also affected by ADHD. For the 493 total individuals within the 73 families, 125 are affected by either ASD or ADHD (Table 1). Among the 98 individuals affected by ASD, 41 (41.8%) are also affected by ADHD (Table 1). Males are more likely to be affected by both ASD and ADHD compared to females (male/female ratio: 2.73, T-test p = 0.03).

**Table 1.** Summary of families and their phenotypes.

|  | All | Male | Female | Male/Female ratio | Families |
| --- | --- | --- | --- | --- | --- |
| ASD | 98 | 77 | 21 | 3.67 | 73 |
| ADHD | 68 | 46 | 22 | 2.09 | 47 |
| ASD and ADHD | 41 | 30 | 11 | 2.73 | 36 |
| ASD or ADHD | 125 | 93 | 32 | 2.91 | 73 |
| All samples | 493 | 281 | 212 | 1.33 | 73 |

### Linkage analysis highlights linkage peaks for ADHD

A total of 524 individuals from 111 families were genotyped for the linkage analysis. The fully constructed pedigrees contain 707 individuals, after adding missing individuals (e.g., grandparents) to link sibships of cousins. Families have an average of 5.7 individuals (including ungenotyped individuals) and 4.2 genotyped individuals.

We performed multipoint linkage analysis of the data for three phenotypes: “ADHD”, where individuals diagnosed with ADHD are considered affected; “ADHD or ASD” (referred to as “ASD/ADHD” in the following text), where both ADHD and ASD affected individuals are considered affected; and “ADHD and ASD”, where only individuals diagnosed with both ADHD and ASD are considered affected. The results are summarized in Figure 2 with peaks summarized in Table S3. The ADHD phenotype was linked to 17p13.1-2, meeting the conventional standard for declaring linkage (PPL=0.38, p<0.001) as determined by a simulation study to estimate the empirical null distribution (Figure 2). Because the pooled PPL was larger than the sequentially updated PPL, we infer that the locus is largely homogenous and these results do not offer evidence that data from either of the two waves are inconsistent with linkage. As such, the ADHD locus did not depend on the strictness of the ASD criteria used in family ascertainment. The locus on 12q12-15 met the criteria for suggestive linkage to ADHD (PPL=0.27, p<0.01, Figure 2). Similar to the locus on 17p, the pooled PPL for 12q was larger than the sequentially updated PPL, indicating that the linkage with ADHD did not differ across the two waves with different ASD recruitment criteria. Several additional loci met the criteria of nominal linkage. The ADHD phenotype is nominally linked to 3p13 (pooled PPL=0.11, sequential PPL=0.23) with evidence for heterogeneity across ASD criteria and 19q12-13.1 (pooled PPL=0.20, sequential PPL=0.14) with no evidence for heterogeneity. The phenotype ADHD/ASD had two nominal linkage peaks, both suggesting heterogeneity, on 19p13.3 (pooled PPL=0.11, sequential PPL=0.18) and 20q13.13-13.33 (pooled PPL=0.02, sequential PPL=0.19). The “ADHD and ASD” phenotype did not show evidence for linkage, likely due to the small sample size of the phenotype (Figure 2).

**Figure 2.**
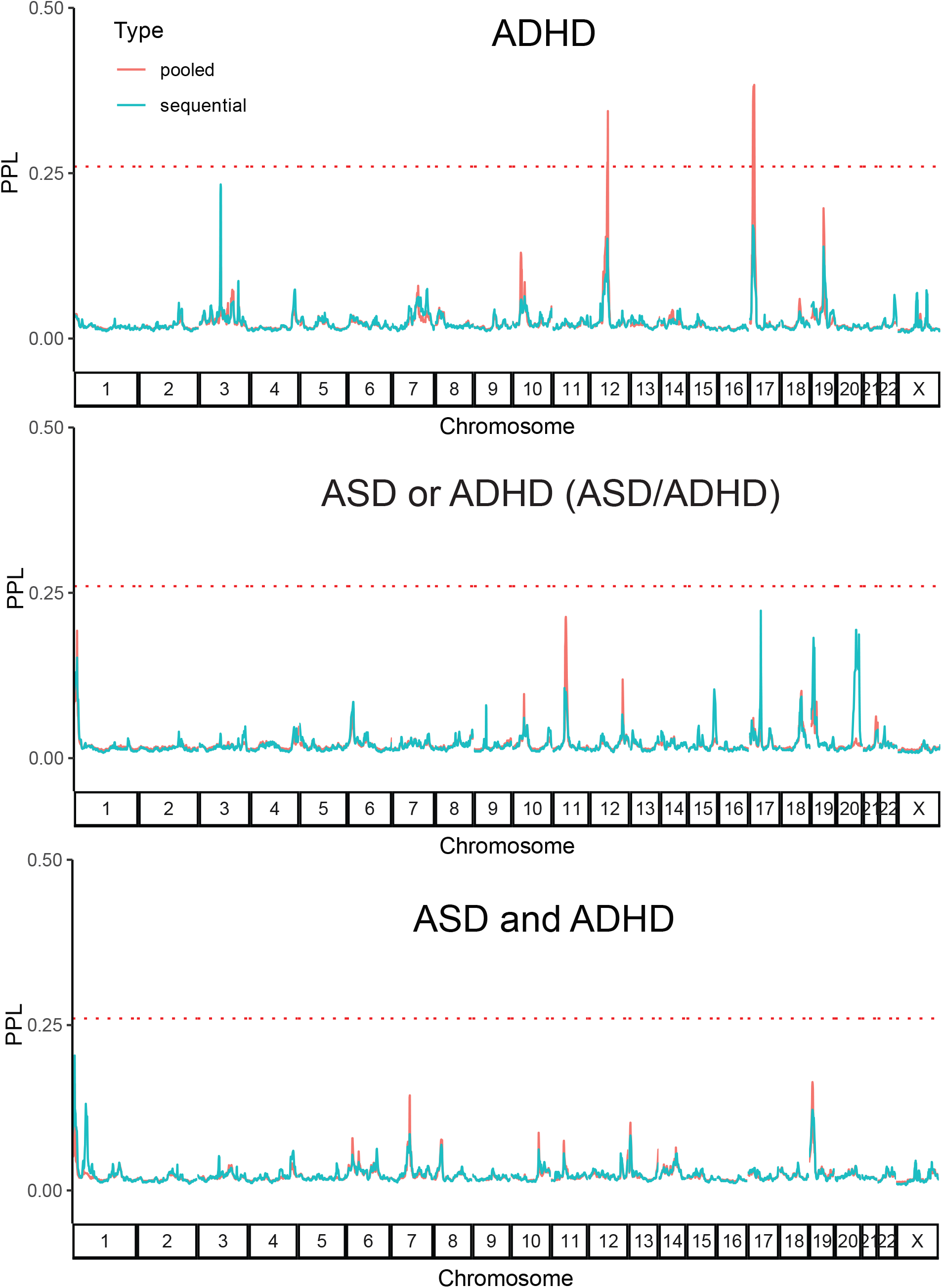
Linkage analysis for ASD and ADHD. Sequential and pooled analysis results are shown in blue and red lines, respectively. The red dotted line indicates the PPL=0.26, which corresponds to p=0.01.

### Whole genome sequencing and variant discovery

We selected 272 individuals from the recruited families to undergo WGS to ∼30x coverage. The sequencing reads that passed quality filters were mapped to the human genome reference hg19 and genomic variants are called jointly using GATK for small variants (SNVs and indels) and MetaSV for SVs.

The number of small variants and SVs were consistent across samples. For small variants, around 4 million SNVs were identified for most of the samples, and the number of indels ranged from 700,000 to 1.2 million (Table S1). For SVs, an average of 743,280 SVs (including unknown type) were discovered for each individual, ranging from 325,443 to 1,291,816 across samples. We selected SVs that were identified by at least two methods for the downstream analysis. An average 1,802 SVs passed the filter in each individual (see Methods for detail), ranging from 1,332 to 2,620 (Table S1). Most SVs are within the 50-1,000 bps range and the largest category is deletion (Figure S1).

### Candidate genes associated with ADHD and ASD/ADHD in linkage regions

To identify the risk genes for ADHD, we first examined SNVs and indels within the chromosome 12 and 17 significant ADHD linkage regions using the pVAAST workflow. pVAAST prioritizes genes by integrating segregation information, population allele frequency, and functional impact of variants in a gene (37). After Bonferroni correction of the p-values (1,299 genes for chromosome 12 and 563 for chromosome 17 linkage regions) and selecting genes express in brain, we identified 34 genes that met the significance criteria (Bonferroni-corrected p<0.05) for the dominant mode and 3 genes for the recessive mode (Table 2). Proline glutamate and leucine rich protein 1 (*PELP1*) is present in both lists. Among the 36 unique genes, 18 are located in the chromosome 12 linkage region and 18 in the chromosome 17 linkage region, respectively. Four genes (*KDM6B, COL2A1, PTPRB, PER1*) were reported in one or more NDD disease gene databases (Table 2). In addition to SNVs and indels, we also identified SVs using the WGS data. Within the linkage regions, an average of 83 SVs were identified per individual, with a minimum of 32 and a maximum of 157 across the cohort. None of the genes passed our SV gene filtering criteria (see whole-genome SV analysis below for filtering detail).

**Table 2.** Candidate genes in linkage regions.

| Gene | Chr | Inheritance | Pvalue | pLI | Database |
| --- | --- | --- | --- | --- | --- |
| *KDM6B* | chr17 | Dominant | 5.63E-04 | 1 | SFARI, iPSYCH |
| *MYH13* | chr17 | Dominant | 5.63E-04 | 0 |  |
| *USP6* | chr17 | Dominant | 5.63E-04 | 0 |  |
| *MYBBP1A* | chr17 | Dominant | 5.63E-04 | 0 |  |
| *KMT2D* | chr12 | Dominant | 1.30E-03 | 1 |  |
| *MYO1A* | chr12 | Dominant | 1.30E-03 | 0 |  |
| *COL2A1* | chr12 | Dominant | 1.30E-03 | 1 | iPSYCH |
| *PTPRB* | chr12 | Dominant | 1.30E-03 | 0.94 | SFARI |
| *ITGA7* | chr12 | Dominant | 1.30E-03 | 0 |  |
| *STAT2* | chr12 | Dominant | 1.30E-03 | 0.04 |  |
| *NEURL4* | chr17 | Dominant | 5.63E-04 | 1 |  |
| *TIMELESS* | chr12 | Dominant | 1.30E-03 | 0 |  |
| *CCDC65* | chr12 | Dominant | 1.30E-03 | 0 |  |
| *PFAS* | chr17 | Dominant | 5.63E-04 | 0 |  |
| *NLRP1* | chr17 | Dominant | 5.63E-04 | 0 |  |
| *EIF4B* | chr12 | Dominant | 1.30E-03 | 1 |  |
| *NELL2* | chr12 | Dominant | 1.30E-03 | 1 |  |
| *WNT10B* | chr12 | Dominant | 1.30E-03 | 0 |  |
| *ALG10B* | chr12 | Dominant | 2.60E-03 | 0 |  |
| *PELP1* | chr17 | Dominant | 1.69E-03 | 1 |  |
| *CHRNE* | chr17 | Dominant | 1.69E-03 | 0 |  |
| *GRASP* | chr12 | Dominant | 3.90E-03 | 0.93 |  |
| *ARHGEF15* | chr17 | Dominant | 2.82E-03 | 0 |  |
| *TROAP* | chr12 | Dominant | 6.50E-03 | 0 |  |
| *TRPV3* | chr17 | Dominant | 4.50E-03 | 0 |  |
| *P2RX5* | chr17 | Dominant | 4.50E-03 | NA |  |
| *DNAJC22* | chr12 | Dominant | 1.04E-02 | 0 |  |
| *ACADVL* | chr17 | Dominant | 1.13E-02 | 0 |  |
| *ZZEF1* | chr17 | Dominant | 1.13E-02 | 1 |  |
| *TEKT1* | chr17 | Dominant | 1.13E-02 | 0 |  |
| *ANKRD33* | chr12 | Dominant | 2.60E-02 | 1 |  |
| *PER1* | chr17 | Dominant | 1.13E-02 | 0.88 | SFARI, ADHDgene |
| *LYZ* | chr12 | Dominant | 2.60E-02 | 0 |  |
| *USP43* | chr17 | Dominant | 1.69E-02 | 0.01 |  |
| *PELP1* | chr17 | Recessive | 5.63E-04 | 1 |  |
| *SHPK* | chr17 | Recessive | 1.13E-03 | 0 |  |
| *ESPL1* | chr12 | Recessive | 3.90E-03 | 1 |  |

The highest-ranking gene is Lysine Demethylase 6B (*KDM6B*) on chromosome 17. *KDM6B* has two missense variants segregating in two families. One mutation (17-7751888-C-G, p.Thr761Ser) is predicted to be possibly damaging by PolyPhen2. The other (17-7749972-G-T, p.Val209Leu) is predicted to be benign by PolyPhen2. Three additional missense variants are also present in patients but do not follow strict segregation patterns (Table S4). *KDM6B* is expressed in the brain and it is intolerant to mutations (pLI=1). The *KDM6B* protein demethylates trimethylated lysine-27 on histone H3. Pathogenic alterations in histone lysine methylation and demethylation genes have been associated with multiple NDDs (47), including ADHD (48, 49). *KDM6B* is also annotated in NDD databases such as SFARI and iPSYCH, and it is identified as one of the 102 risk genes in a recent large ASD study (50).

The highest-ranking gene in the chromosome 12 linkage region is lysine methyltransferase 2D (*KMT2D*), which is also involved in lysine methylation, similar to *KDM6B*. Two missense variants segregate in two families (12-49428694-T-C, p.Asp3419Gly; 12-49436073-C-T, p.Asp1970Asn), both are predicted to be damaging by both SIFT and PolyPhen2. Nine additional missense variants are present in patients but do not follow strict segregation patterns (Table S4). *KMT2D* is expressed in the brain and intolerant to mutations (pLI=1).

Another candidate gene from the chromosome 17 linkage region, Neuralized E3 Ubiquitin Protein Ligase 4 (*NEURL4*), has a missense variant (17-7224433-C-T, p.Gly1120Arg) segregating in one family and four additional missense variants in other affected individuals. *NEURL4* is expressed in the brain and intolerant to mutations (pLI∼1). *NEURL4* protein is a scaffold protein, which maintains normal centriolar homeostasis and prevents formation of ectopic microtubule organizing centers (51). Although no publication directly links *NEURL4* with NDDs, evidence has shown that microtubule related genes are related to neuronal migration and dendritic functioning (52). Knockout of *NEURL4* in mice causes neurological phenotypes such as decreased prepulse inhibition (43).

In the chromosome 12 linkage region, we also identified the circadian rhythm controller gene Timeless Circadian Regulator (*TIMELESS*) with a missense mutation (12-56827209-C-A, p.Ala129Ser). The mutation is segregated in two families and is predicted to be deleterious by SIFT, suggesting its potential role in ADHD etiology. Sleep disorders are common in both ASD and ADHD patients (53). A previous study found mutations in circadian-relevant genes, including *TIMELESS*, are more frequent in ASD affected individuals than controls (54).

### Novel and known risk genes associated with ADHD and ASD/ADHD in the whole genome analysis

Because ASD and ADHD are highly heterogeneous and multiple variants/genes could contribute to their genetic etiology, next we performed pVAAST analysis on SNVs/indels for the whole genome. After selecting genes with evidence of inheritance (LOD score>=1) and express in the brain (see Methods for detail), we identified 16, 25, 70, and 29 candidate genes for ADHD dominant model, ADHD recessive model, ASD/ADHD dominant model, and ASD/ADHD recessive model, respectively (Table S5).

From SVs, we identified 209 and 227 genes that contain exonic SVs in at least one affected individual for ADHD and ASD/ADHD, respectively (Table S6). To further prioritize genes affected by SVs, we calculated the combined severity scores of SVs within each gene and determined their brain expression levels. We then selected genes whose combined severity scores were larger in affected individuals than in unaffected ones and were expressed in the brain. A total of 43 and 44 genes passed our SV gene filtering criteria for ADHD and ASD/ADHD, respectively (Table S6).

Next, we created two high-confidence candidate gene sets for ADHD and ASD/ADHD. For ADHD, we combined significant linkage-region candidate genes, high-confidence candidate genes from the whole-genome pVAAST analyses, and the SV analysis. For ASD/ADHD, we combined candidate genes from whole-genome pVAAST and SV analyses. A total of 207 unique genes were included in these two confidence gene sets, including 113 genes for ADHD and 138 for ASD/ADHD (Table 3). We annotated these genes using public resources, including gene association databases, literature search databases, tolerance to loss of function mutation scores, mouse knock-out experiments, and expression databases (Table S7, see Methods for detail). A total of 31 genes (15%) were described by previous studies or existing NDD gene databases (Table S2), including 22 genes from the SFARI database. Three genes (*ANKRD11, KDM6B, COL2A1*) overlap risk genes in the iPSYCH study of ASD and ADHD (55).

**Table 3.** Candidate gene summary.

|  | SNV/Indel |  | SV |  | Total<br>(unique) |  |
| --- | --- | --- | --- | --- | --- | --- |
|  | Genes | Families* | Genes | Families* | Genes | Families* |
| **ADHD** |  |  |  |  |  |  |
| Linkage | 36 | 19 | 0 | 0 | 36 | 19 |
| WGS | 40 | 47 | 43 | 42 | 83 | 47 |
| Total (Unique) | 70 | 47 | 43 | 42 | 113 | 47 |
| **ASD/ADHD** |  |  |  |  |  |  |
| WGS | 94 | 73 | 44 | 73 | 138 | 73 |
| **Total (Unique)** | 140 | 73 | 61 | 73 | 207 | 73 |

\*For SNV/indel, families with segregating variants in a given gene were counted. For SV, families with at least one affected individual with an exonic SV were counted

In addition to genes overlapping known NDD genes, many other candidate genes showed functional relevance to neurodevelopmental processes or have been reported to be associated with NDDs. For example, we identified Protein disulfide-isomerase A2 (*PDIA2*) in the whole-genome pVAAST analysis. With the ADHD dominant model, *PDIA2* had a missense mutation (16-334899-G-A, p.Ala188Thr) segregating in two families and was predicted as damaging by SIFT. It had additional 27 variants segregating in 13 families. *PDIA2* encodes a member of disulfide isomerase family of endoplasmic reticulum (ER) proteins that catalyze protein folding and thiol-disulfide interchange reactions and is highly expressed in the cerebellar cortex. HumanBase predicted a strong association of *PDIA2* to ASD (rank 1) and a study investigating the role of ER stress in autism identified *PDIA2* upregulation in autistic subjects (56).

In whole-genome SV analysis, we discovered a novel candidate gene Calcium Voltage-Gated Channel Auxiliary Subunit Alpha2delta 4 (*CACNA2D4*). *CACNA2D4* is one of the genes that encode the alpha-2/delta subunit of the voltage-dependent calcium channel, among three other genes (*CACNA2D1, CACNA2D2, CACNA2D3*) (57). While a dozen genes encoding the alpha1 and beta subunits of the calcium channel are previously identified in the SFARI database (Table S2), *CACNA2D4* was absent. In our study, *CACNA2D4* was found to contain SVs in affected individuals in five families in the ASD/ADHD analysis (one SV overlaps intron 16 to intron 26 and other SVs within intron 26). The SV overlapping intron 16 to intron 26 is a 36 kb deletion (chr12:1948911-1985036) that is rare in general populations (<0.16% in European populations in gnomAD, https://gnomad.broadinstitute.org/variant/DEL_12_124891?dataset=gnomad_sv_r2_1). Two previous ASD studies identified *CACNA2D4* deletions in ASD patients and support our finding (58, 59).

### Candidate genes participate in pathways related to neurological disorders

To explore relationships among the candidate genes and to identify potential shared mechanisms between ASD, ADHD, and other NDDs, we performed enrichment analysis for the 207 candidate genes with 1,629 known NDD genes (Table S2) and determined the significance of enrichment for each gene list (Table S8). ADHD and ASD/ADHD showed strong enrichment in GO terms and pathways related to cilium and microtubule, suggesting that many genes in this group are involved in brain development (Table S8). For example, pathway “Ciliopathy” (WP4803) is significantly enriched in both ADHD and ASD/ADHD gene sets, overlapping five and seven genes in the two sets, respectively. “cilium or flagellum-dependent cell motility” (GO:0001539) is one of the top enriched GO term in ADHD gene list, and overlap four genes in the candidate genes (*BBS4, CCDC65, TEKT1, TEKT5*).

Other processes that are known to relate to NDDs are also present. For example, “cation channel activity” (GO:0005261) is enriched in both ADHD and ASD/ADHD gene sets, overlapping seven and eight genes in the two sets, respectively. Besides shared enriched terms and pathways, each gene set also contains phenotype-specific enrichments. For example, striatum development (GO:0021756) and cerebral cortex development (GO:0021987) are significantly enriched in the ASD/ADHD gene set, overlapping two and four candidate genes, respectively. On the other hand, “sensory perception of light stimulus” is only significantly enriched in ADHD gene set (GO:0050953), overlapping seven genes in the set.

### Protein-protein interaction network of candidate genes

To further examine the relationship among the candidate genes, we constructed PPI networks using our candidate genes and known NDD genes (Figure 3, Table S9). Of the 113 ADHD genes, 23 were connected with known NDD genes into a single PPI network with interactions supported by at least 2 databases (Figure 3A, 45 genes in total), and 50 of the remaining genes were connected with known NDD genes with interactions supported by at least one database (Figure 3B, 150 genes). Similarly, for the 138 ASD/ADHD genes, 44 were connected after adding NDD genes with interactions supported by ≥ 2 databases (Figure 3C, 71 genes in total), and 31 of the remaining genes were connected after adding NDD genes with interactions in ≥ 1 database (Figure 3D, 83 genes). Several enriched terms, such as histone methyltransferase complex and cation channel activity, showed extensive connection with known genes for ASD, ADHD, and other NDDs, supporting the shared genetic etiology of ADHD and ASD.

**Figure 3.**
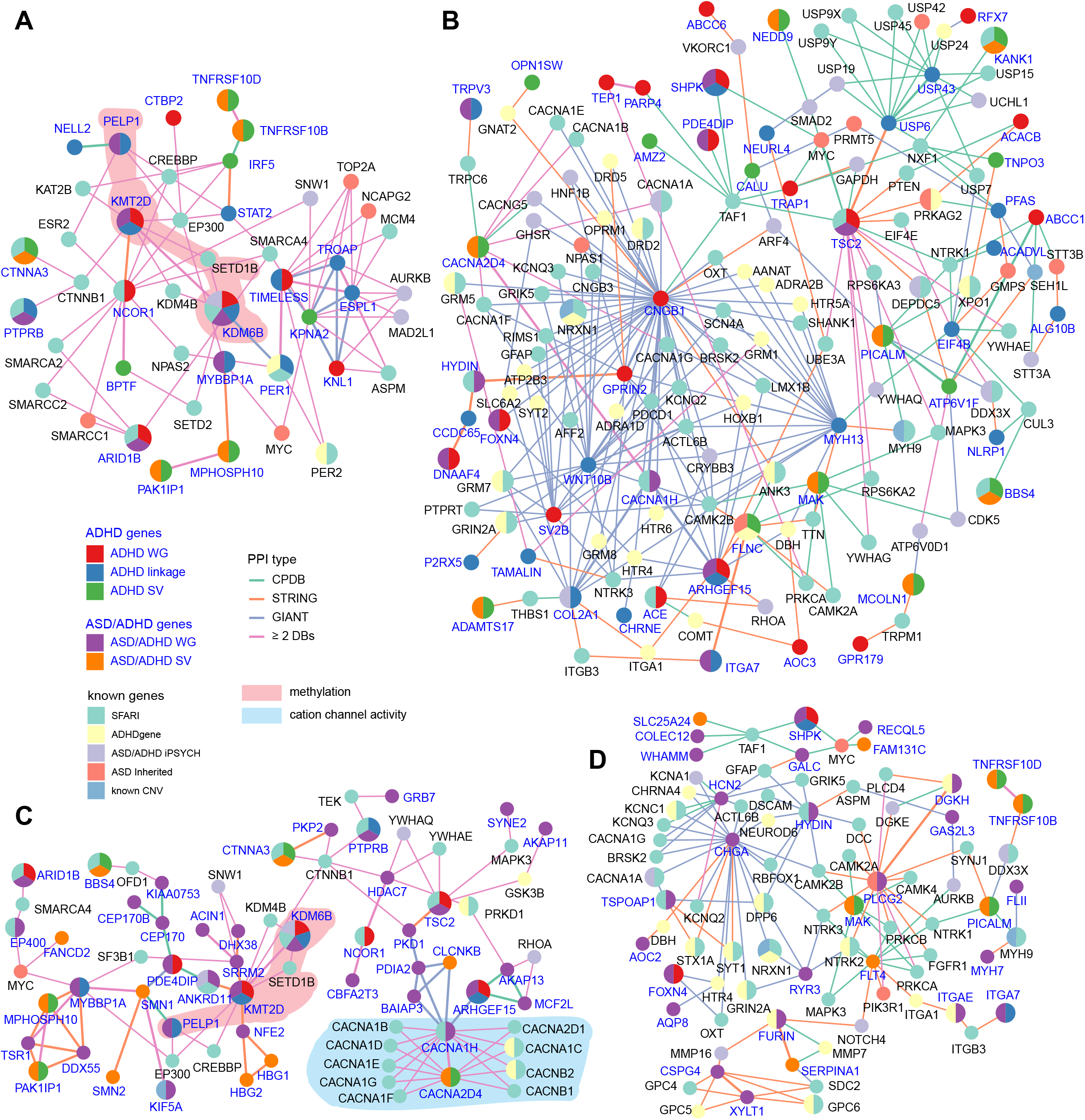
Protein-protein interaction (PPI) networks of candidate genes. Candidate genes were ADHD risk genes in (**A, B**) and ASD/ADHD risk genes in (**C, D**). (**A, C**): PPI networks of the candidate genes. Other known NDD genes were added as intermediate nodes if they interact with more than one candidate gene supported by PPI evidence from ≥ 2 databases. (**B, D**): PPI network of candidate genes that are not in (**A**) and (**C**), respectively. Known genes were added as intermediate nodes if they interact with more than one candidate gene with PPI evidence from any PPI database. PPI networks were defined by three databases, ConsensusPathDB, STRING, and GIANT_v2. Genes were colored by gene lists. ASD/ADHD genes were used to color genes in (**A, B**) and ADHD genes were used to color genes in (**C, D**). To simplify the network, PPIs between non-candidate genes were removed. Genes belonging to two GO terms, methylation and cation channel activity, were highlighted in **A, C**, respectively.

## Discussion

ASD and ADHD co-occur at a high frequency and studies suggested shared genetic risk factors between the two disorders (1–7). Given the worsened negative impact on patients’ quality of life when the two conditions co-occur, it is crucial to understand the underlying genetic risk factors for each disorder and the co-occurrence. In the NJLAGS cohort, ADHD occurs in 41.8% of ASD patients, consistent with previous studies (reviewed in (60)). The family design of the NJLAGS study provides a unique opportunity to identify inherent and *de novo* genetic factors underlying both ASD and ADHD.

In this study, we identified two linkage peaks (12q12-15, 17p13.1-2) for the ADHD phenotype from 111 families. Using WGS data from 272 selected samples, we discovered 36 significant genes within the linkage regions and explored their gene functions. Expanding the analysis to the whole genome revealed more risk genes for both ADHD and ASD/ADHD phenotypes. Notably, both top candidate genes in the two linkage regions (*KDM6B, KMT2D*) are involved in the histone lysine methylation process. Histone modification is a central process regulating gene expression during neurodevelopment and mutations in *KDM6B* have been linked to multiple NDDs (47), including ADHD (48, 49). At the whole genome level, 31 of the high-confidence candidate genes were identified in previous studies or NDD gene databases. In addition, we observed multiple NDD-related pathways overrepresented in the high-confidence ADHD and ASD/ADHD gene sets (Table S8). Taken together, the results demonstrate the power of combining linkage analysis, rare variant analysis, and SV analyses to uncover risk genes.

By studying ADHD affected patients in families with ASD probands, we identified genetic risk factors underlying ASD and ADHD co-occurrence. Three candidate genes (*ANKRD11, KDM6B, COL2A1*) were reported in a previous study of ASD and ADHD comorbidity in Danish cohorts (55). In the Danish cohorts, all three genes contain multiple protein-truncating variants or missense variants in ASD or ADHD patients, and *ANKRD11* (Ankyrin Repeat Domain Containing 11) is one of the top candidate genes with a significant excess of constrained rare protein-truncating variants (crPTVs) (55). In addition, several genes (*KDM6B, PTPRB, PER1*) that are significant within the ADHD linkage region are described in previous ASD studies (61–63). Pathways we discovered that are important in ADHD etiology are also vital in ASD etiology. By providing an extensive candidate gene set, we are not only reaffirming the linkage between ASD and ADHD, but also suggesting novel potential risk pathways for further investigation.

Several important GO terms and pathways recurred in both gene sets in our enrichment analysis, namely cilium/microtubule, ion channel, histone methylation, and chromatin remodeling. These pathways revealed interesting aspects of mechanisms in ASD and ADHD etiology as an interconnected network of hundreds of genes and pathways (64) and underlined the complexity of genetic casualties of NDDs. A recent article argued that several NDDs share common genetic risk factors and there might not be “Autism-specific” genes (65). Our results support this view. We also identified some important but underappreciated processes such as the circadian rhythm (GO:0007623), which has phenotypic effects in cognition, mood, and reward-related behaviors. Research documented that the prevalence of insomnia ranges from 50% to 80% in ASD patients, compared to 9–50% in age-matched typically developing children (66). However, the underlying mechanism of association and genes involved remain to be determined. The genes related to circadian rhythm we identified in this study (*MYBBP1A, PER1, TIMELESS*) could shed light on the genetic etiology of sleep disorders in ASD and ADHD patients.

Our study has a few limitations. The limited sequencing capacity led to the sequencing of only a selected number of ADHD affected individuals, as the NJLAGS dataset was collected primarily to identify ASD and language-impaired probands. However, recently we sent out follow-up questionnaires to all NJLAGS families requesting an update on ADHD diagnoses. We specifically asked who had received a diagnosis from a medical professional (*i*.*e*., psychiatrist, developmental pediatrician, neurologist, psychologist). Of the 34 families who responded to our questionnaire, 15 were families who had some sequencing data and whose data are included in this manuscript. From these 15 families, 28 individuals had received a formal diagnosis of ADD/ADHD by a medical professional. Importantly, only 13 of these family members had been initially identified as having a diagnosis of ADD/ADHD. In some cases these were younger family members who were too young at the time of data collection to be identified as affected while in other cases, we did not have this diagnostic information available. These updated findings will be included in future ADHD in genetic analyses. Moving forward, we anticipate extensive sequencing efforts to continue for the NJLAGS project resulting in the sequencing of all NJLAGS families. As more ADHD affected individuals are identified and sequenced, this more complete pedigree information and stronger statistical power will allow us to further discover potential common risk factors underlying the ASD and ADHD etiology.

Another limitation of this study is we focused mainly on variants affecting protein coding sequences. Non-coding regulatory variants are known to play an important role in NDDs, although predicting their functional impact is still difficult (67). With the rapid development of functional prediction algorithms for non-coding variants, we will identify additional candidate genes affected by regulatory variants using the cohort’s WGS data in the near future.

In conclusion, using linkage analysis and WGS-based variant analysis, we examined the genetic etiology of ASD and ADHD in the NJLAGS cohort. The family-based design of the cohort allows us to leverage the power of variant segregation while taking into account the phenotype heterogeneity of ASD and ADHD. By examining both small variants and large SVs, we identified genes and pathways, previously known or novel, that could contribute to ASD or ADHD etiology. Because of the difficulty of distinguishing some of the ASD and ADHD behavioral characteristics, understanding the genetic risk factors for the disorders can provide guidance to clinical intervention approaches, both behaviorally and pharmacologically. Future studies, especially functional assessment of these candidate genes, will elucidate their roles in ASD and ADHD and improve the diagnosis and treatment of these disorders.

## Data Availability

The raw sequencing reads, variants, and genotypes for all samples are available online at the National Institute of Mental Health Data Archive (NDA) under experiments C1932 and C2933.

https://nda.nih.gov/

## Acknowledgments

We thank the patients who participated in and contributed to this study. We gratefully acknowledge access to the HPC facilities and support of the computational STEM and bioinformatics scientists from the Office of Advanced Research Computing at Rutgers University. This study was supported by NIMH grants R01 MH070366 and RC1 MH088288 to LMB; New Jersey Governor’s Council for Medical Research and Treatment of Autism grant CAUT12APS006 to LMB and CAUT19APL028 to JX and LMB.

## Supplementary information

Supplementary information is available at MP’s website

