## Supplementary material for "Common Genetic Risk Factors in ASD and ADHD Co-occurring Families": Figure S1

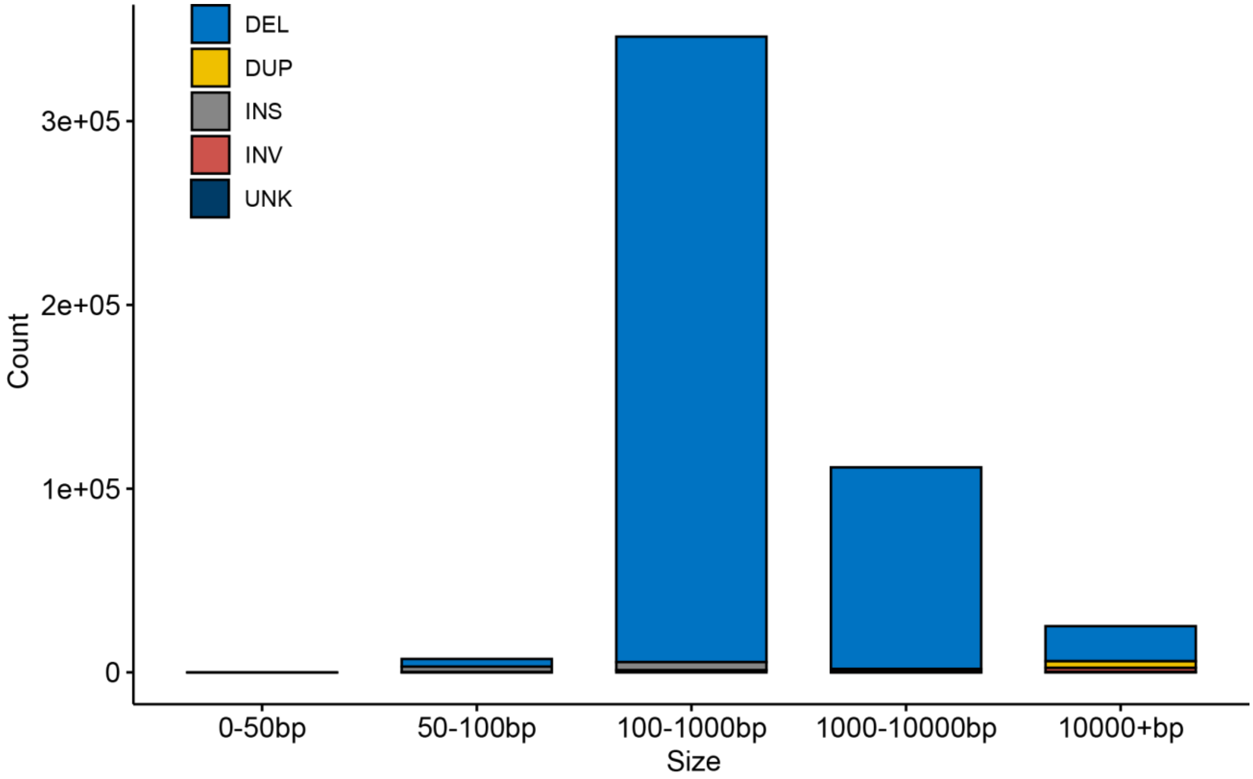


**Figure S1. Size distribution of SVs.** SV types: DEL: deletion; DUP: duplication; INS: insertion; INV: inversion, UNK: other types, including interspersed duplication (IDP) and intra-chromosomal translocation (ITX).
